# Stop LCNP: High dose corticosteroid therapy for late radiation-associated lower cranial neuropathy: A report of the phase I dose finding trial and parallel prospective data registry

**DOI:** 10.64898/2026.06.17.26354728

**Authors:** Zayne A.S. Belal, Christine B. Peterson, Carly E. Barbon, Holly McMillan, Sheila N. Buoy, Jose A. Garcia, Nicolaas C. Anderson, Clifton D. Fuller, Karin Woodman, Stephen Y. Lai, Katherine A. Hutcheson

**Affiliations:** 1Department of Radiation Oncology, University of Pennsylvania, Philadelphia, PA; 2Department of Biostatistics, The University of Texas MD Anderson Cancer Center, Houston, TX; 3Department of Head and Neck Surgery, The University of Texas MD Anderson Cancer Center, Houston, TX; 4Department of Neurology, Baylor College of Medicine, Houston, TX; 5Department of Radiation Oncology, The University of Texas MD Anderson Cancer Center, Houston, TX; 6Department of Neurology, Vanderbilt University Medical Center, Nashville, TN

**Author notes:** Denotes first Author (Zayne A.S. Belal). Denotes Corresponding Author **Corresponding Author Name & Email Address:** Kate A. Hutcheson, Stephen Y. Lai, and Karin Woodman,.

**Keywords:** Radiation-associated lower cranial neuropathy, Corticosteroid therapy, Head and neck cancer survivorship, Dysphagia, Phase I clinical trial, Late radiation toxicity, Symptom burden, MDASI-HN

## Abstract

**Purpose:** Radiation-associated lower cranial neuropathy (LCNP) is a debilitating late complication among head and neck cancer (HNC) survivors, leading to progressive dysphagia, aspiration, and loss of nutritional independence. No proven therapies exist to reverse LCNP. This Phase I dose-finding trial (XXXX, XXXX) and parallel registry study prospectively evaluated the safety, feasibility, tolerability, and symptomatic response of high-dose corticosteroid therapy for radiation-associated LCNP.

**Methods and Materials:** Eligible participants were disease-free oropharyngeal cancer survivors ≥2 years post-radiotherapy with LCNP involving CN XII ± X and no structural or malignant etiology. Phase I trial participants received oral prednisone 1 mg/kg (Dose 1, n = 3) or 3 mg/kg (Dose 2, n = 5) daily for 5 days followed by a 2-week taper. A parallel registry (n = 6) enrolled broader HNC survivors treated with 1 mg/kg. Phase I dose escalation decisions were based on the balance of tolerability and symptom response. The primary endpoint was change in MDASI-HN Top 5 mean symptom score from baseline to 1–2 weeks post-taper; a ≥ 1.315 unit decrease indicated clinically meaningful improvement. Secondary endpoints included clinician-graded measures of bulbar function, electromyography (EMG) and patient-report outcome measures.

**Results:** All regimens were feasible and well tolerated. Insomnia (n = 4, Grade 1) was the most frequent adverse event and there were no treatment discontinuations. At 1–2 weeks post-taper, the median MDASI-HN Top 5 change was −0.2 in Dose 1 and −1.6 in Dose 2; three of five Dose 2 patients achieved clinically meaningful improvement versus one in Dose 1. By 6–10 weeks, symptom improvement persisted in a subset but diminished overall. Dose 2 met pre-specified criteria for Phase II progression. No consistent improvements were observed in objective functional or electrophysiologic measures, though exploratory trends favored higher dosing.

**Conclusions:** High-dose corticosteroid therapy at 3 mg/kg/day was feasible and tolerable in long-term HNC survivors with LCNP and showed preliminary evidence of short-term symptomatic benefit. Phase II evaluation is warranted.

## Introduction

The epidemiology of head and neck cancer (HNC) has evolved significantly, largely driven by the increasing incidence of human papillomavirus (HPV)-associated oropharyngeal cancer (OPC), which is projected to continue rising over the next two decades [1–7]. Concurrently, improvements in treatment strategies, particularly the widespread adoption of organ-preserving radiotherapy (RT) and chemoradiotherapy, have led to steadily increasing survival rates, with a growing proportion of long-term survivors now cured of their disease. However, this progress has also resulted in a rising prevalence of late radiation-associated toxicities, motivating increased attention towards mitigating long-term treatment effects in this population [8].

One of the most disabling late effects in HNC survivors is radiation-associated lower cranial neuropathy (LCNP), a rare but severe complication that can emerge years after treatment. Reports estimate that 5% of OPC survivors develop LCNP by five years post-treatment, with a cumulative lifetime risk exceeding 10% [9]. LCNP primarily affects cranial nerves (CN) IX, X, and XII, leading to progressive bulbar dysfunction that manifests as dysphagia, aspiration, dysphonia, and dysarthria. Prior research among nearly 900 OPC survivors demonstrated significantly higher symptom burden in those with LCNP, with the most pronounced impact on swallowing and speech [10]. Case studies further highlight the profound functional impairments and diminished quality of life (QOL) associated with LCNP [11,12].

Currently, there are no proven therapies to reverse LCNP, leaving most survivors who develop cranial neuropathy with severe swallowing dysfunction at high risk for aspiration pneumonia, chronic feeding tube dependence, and increased mortality. In a prior series, 67% of OPC survivors with late LCNP-associated dysphagia required lifelong feeding tube dependence, typically occurring at a median age of 64 years, nearly a decade after cancer cure [13]. The physical, social, and emotional consequences of such disability at an active stage of life are profound.

Case reports suggest that high-dose corticosteroid therapy may offer a promising intervention for radiation-associated LCNP. Two published cases demonstrated functional reinnervation on electromyography (EMG) and improved swallowing function on modified barium swallow (MBS) studies following corticosteroid treatment [14,15]; one tested oral prednisone and the other intravenous solumedrol High-dose corticosteroids are routinely used for radiation-induced plexopathy, acute multiple sclerosis exacerbations, and other acquired neuropathic conditions, such as chronic inflammatory demyelinating polyneuropathy [16]. However, no clinical trials have systematically evaluated the feasibility, optimal dosing, or functional impact of corticosteroid therapy for radiation-associated LCNP in HNC survivors.

### Study rationale and objectives

Given the profound morbidity associated with LCNP and the lack of established treatment options, we designed a Phase I/II dose-finding clinical trial to evaluate corticosteroid therapy for this unique indication. The primary objective was to determine the feasibility and maximum tolerated dose (MTD) of corticosteroid therapy that provides symptomatic improvement in OPC survivors with radiation-associated LCNP. The secondary objective was to assess potential functional and symptomatic gains, including changes in swallowing function, speech, QOL, neurophysiology, and imaging outcomes following corticosteroid treatment. If successful, this study would provide critical evidence supporting the development of a therapeutic strategy for a previously refractory late radiation toxicity in HNC survivors.

## Methods and Materials

### Study design

XXXX is a prospective, single-institution Phase I/II dose-finding trial (NCT_XXXX) conducted at XXXX (IRB #XXXX). The dose escalation schema is shown in Figure 1 and the CONSORT flow diagram for this trial is shown in Supplementary Figure 1. This report summarizes results from the Phase I dose escalation (Dose 1 cohort 1 and Dose 2 cohort 1); the intravenous dose expansion phase (Dose 3) was not initiated yet. In Phase I each dose level enrolled 3–5 patients to assess safety and symptomatic response. If Phase I criteria for tolerability and symptom reduction were achieved, Phase II accrual would continue at that dose up to 15 total patients to better characterize achievable effect sizes and adverse event (AE) profiles.

**Figure 1:**
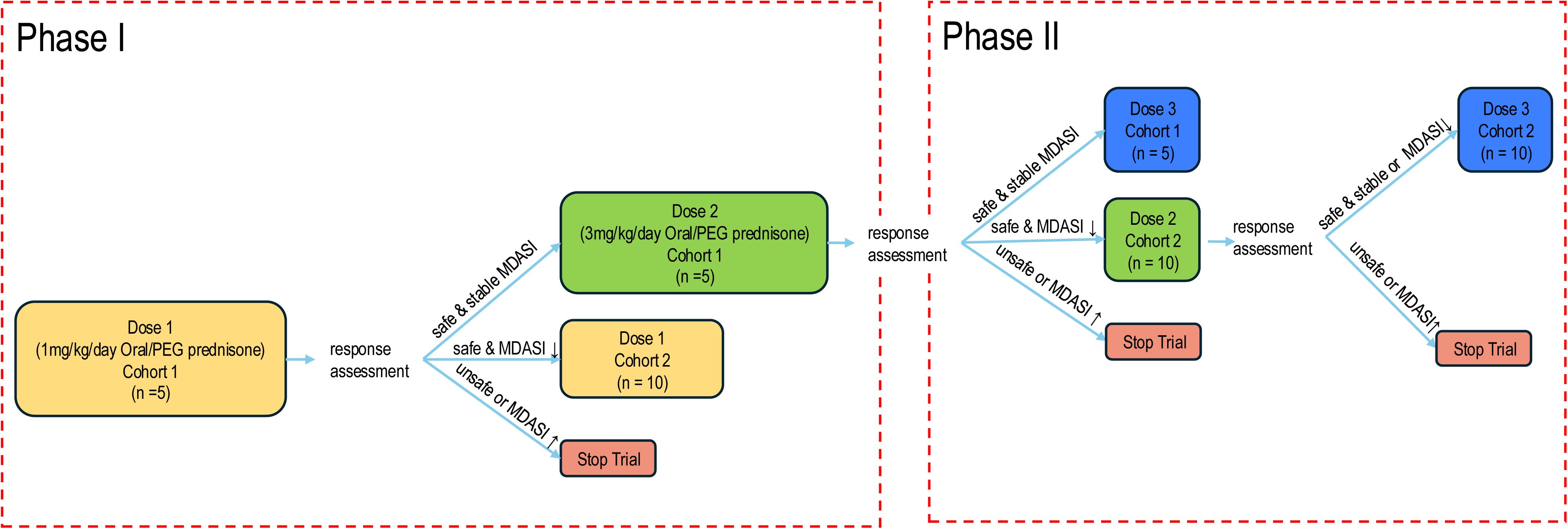
XXXX study schema.

A parallel registry study to the Phase I/II trial allowed for prospective data collection among participants treated with high dose corticosteroid therapy for treatment related LCNP who were ineligible for the Phase I/II trial. All participants were provided written informed consent before any study-specific procedures were performed.

### Patient selection and eligibility criteria

Eligibility criteria is summarized in Supplementary Table 1 included disease-free adult OPC survivors ≥2 years post-radiotherapy with radiation-associated LCNP affecting CN XII ± CN X and no structural or malignant cause identified. Other key exclusion criteria are summarized in Supplementary Table 1. The registry cohort had broader inclusion, allowing any head and neck cancer type.

### Steroid administration and dose selection

Steroids were administered orally or via gastrostomy tube. Dose 1 and registry patients received 1 mg/kg of oral prednisone (±10%) daily for 5 days, followed by a 2-week prednisone taper (±4 days). This represents the lowest reported dose for this indication in the literature [15]. Dose 2 patients received 3 mg/kg of oral prednisone (±10%) daily for 5 days, followed by the same tapering schedule using a 50% dose reduction every 3 days or a more gradual taper at the treating physician’s discretion. This dose was pragmatically selected by the trial neurologist as a clinically meaningful escalation expected to be tolerable and potentially more effective.

Dose 3 (not tested in Phase I) was predefined as 1000 mg of intravenous methylprednisolone daily for 5 days, followed by a 2-week prednisone taper. This dose is used for autoimmune neuromuscular complications and is standard in the management of multiple sclerosis exacerbations [16–23]. A 5-day regimen was favored over weekly 24-hour infusions commonly used for spinal cord injury due to feasibility concerns for long-term survivors with travel limitations.

### Assessments and outcome measures

Study assessments were conducted at baseline (pre-steroid), 1–2 weeks post-taper (1–2w), and 6–10 weeks post-taper (6–10w), with only the **** Symptom Inventory–Head and Neck Module (MDASI-HN) collected longitudinally through 2 years post-treatment. Data were captured using REDCap (v15.2.0; ****), a secure, web-based platform hosted at **** [24,25]. Supplementary Table 2 summarizes all study instruments and assessment schedules, including clinical studies such as tongue electromyography (EMG), modified barium swallow (MBS), and objective lingual function measures, as well as validated questionnaires MDADI and PSS-HN. These were collected at the three primary timepoints but not longitudinally through 2 years of follow-up.

### Primary and secondary endpoints

The primary response endpoint was the MDASI-HN top 5 mean score, defined as the average of the five most severe symptom items among the 22 MDASI-HN symptoms. The five symptoms assessed were difficulty swallowing/chewing, choking/coughing when swallowing, dry mouth, thick mucus, and problems with voice/speech, based on the most commonly reported symptoms in a prior cohort of long-term LCNP survivors at our institution [10]. Symptom response was classified based on change from baseline at each timepoint and improvement was defined as a decrease of ≥0.5 standard deviations, stability as a change of <0.5 SD, and worsening as an increase of ≥0.5 SD, based on prior data in LCNP survivors where 0.5 SD = 1.315 units [10].

Electrophysiologic evaluation was conducted by the trial neurologist as a screening step and included EMG and nerve conduction studies. Denervation was graded using the Muscle Grading (MG) scale (0–4), while reinnervation was assessed via a 1–4 scale based on motor unit action potential (MUAP) characteristics [26,27,28]. Changes in genioglossus denervation and reinnervation scores were classified as improved (bilateral improvement), stable (no change bilaterally), mixed (discordant unilateral changes), or worse (bilateral worsening). Worsening was defined as an increase in denervation or reinnervation score severity relative to baseline.

Secondary patient-reported outcome (PRO) and clinician-graded endpoints are described in Supplementary Table 2 [29,30,31,32,33,34].

### Feasibility and tolerability criteria

The treatment regimen was deemed feasible if ≥80% of participants completed the prescribed regimen and tolerable if ≤2 patients per cohort discontinued corticosteroids or experienced Grade ≥3 toxicity. Adverse events were assessed at each study visit using the Common Terminology Criteria for Adverse Events (CTCAE v5.0) and summarized descriptively. If ≥3 subjects tolerated and demonstrated symptom improvement (per top 5 MDASI-HN change) in Phase I, the trial proceeded to Phase II at the same dose level.

### Statistical analysis

Assessment scores and outcome measures were exported from REDCap (version 15.2.0) [24,25] and processed in Microsoft Excel (Microsoft Corporation, Redmond, WA), JMP (version 18, SAS Institute Inc., Cary, NC), and Python (version 3.13, Python Software Foundation, Wilmington, DE). Descriptive statistics were used to summarize patient-reported outcomes (MDASI-HN top 5), EMG-based denervation/reinnervation scores, DIGEST and PSS-HN swallowing assessments, and lingual function measures. To evaluate symptom response within each dose level, a paired t-test was used to compare MDASI-HN top 5 scores at baseline versus 1–2 weeks post-taper. The change in symptom burden (ΔMDASI-HN top 5) was calculated for each patient, and a one-sample t-test was performed on these deltas against a null hypothesis of no change (mean = 0). Analyses were stratified by dose level. Data visualization was performed using R (version 4.3.1, R Foundation for Statistical Computing, Vienna, Austria), Excel, and GraphPad Prism (version 10.3.1, GraphPad Software, San Diego, CA).

## Results

### Study demographics

A total of 14 patients were enrolled across three groups: Phase I Dose 1 (1 mg/kg, n=3), Dose 2 (3 mg/kg, n=5), and the Registry group (1 mg/kg, n=6). Most were age 55-64 with an equal number of males and females. Full demographic and disease characteristics are detailed in Table 1.

**Table 1:** Demographic and clinical characteristics of the patients enrolled in Phase I of the XXXX clinical trial, and parallel registry study.

|  | Phase I Trial Dose 1<br>(n=3) | Phase I Trial Dose 2<br>(n=5) | Prospective Registry<br>(n=6) |
| --- | --- | --- | --- |
| **Age (years)** |  |  |  |
| <55 | 0 | 0 | 2 |
| 55-64 | 1 | 4 | 3 |
| 65-74 | 1 | 1 | 0 |
| 75-85 | 1 | 0 | 1 |
| **Gender** |  |  |  |
| Male | 0 | 1 | 6 |
| Female | 3 | 4 | 0 |
| **Disease site** |  |  |  |
| Unknown Primary | 1 | 0 | 0 |
| Oropharynx | 2 | 5 | 2 |
| Nasopharynx | 0 | 0 | 3 |
| Other | 0 | 0 | 1 |
| **Disease subsite** |  |  |  |
| Tonsil | 0 | 2 | 1 |
| Base of Tongue | 2 | 3 | 1 |
| Nasopharynx | 0 | 0 | 3 |
| Unknown Primary | 1 | 0 | 0 |
| Other | 0 | 0 | 1 |
| **T-staging\*** |  |  |  |
| 0 | 1 | 1 | 0 |
| 1 | 0 | 0 | 1 |
| 2 | 1 | 4 | 3 |
| 3 | 1 | 0 | 0 |
| 4 | 0 | 0 | 2 |
| **N-staging\*** |  |  |  |
| 0 | 0 | 2 | 1 |
| 1 | 0 | 0 | 1 |
| 2 | 0 | 1 | 1 |
| 2a | 0 | 0 | 0 |
| 2b | 2 | 0 | 1 |
| 2c | 1 | 1 | 2 |
| 3 | 0 | 1 | 0 |
| **M-staging\*** |  |  |  |
| 0 | 3 | 5 | 6 |
| **Prescribed RT Dose to Primary (Gy)** |  |  |  |
| Unknown | 0 | 1 | 1 |
| 66-70 | 3 | 4 | 5 |
| **Chemotherapy** |  |  |  |
| No Chemotherapy | 0 | 1 | 1 |
| Concurrent | 1 | 2 | 3 |
| Induction | 2 | 0 | 1 |
| Induction + Concurrent | 0 | 2 | 1 |
| **Surgery** |  |  |  |
| No surgery | 2 | 4 | 3 |
| Primary Site | 0 | 0 | 0 |
| Neck Dissection | 1 | 1 | 0 |
| Primary site + Neck Dissection | 0 | 0 | 3 |
| Affected Nerve** |  |  |  |
| CN IX & X | 1 | 4 | 3 |
| CN XI | 2 | 5 | 5 |
| CN XII | 3 | 5 | 5 |
| CN XII LCNP Laterality*** |  |  |  |
| Ipsilateral | 0 | 3 | 3 |
| Contralateral | 0 | 0 | 0 |
| Bilateral | 3 | 2 | 2 |
| Abbreviations: LCNP = Lower Cranial Neuropathy, RT = Radiotherapy in units of Gray (Gy), Concurrent = Concurrent Chemoradiotherapy, CN = Cranial Nerve<br>Data in table presented as absolute counts (n)<br>*TNM Staging based on AJCC 7<sup>th</sup> edition staging for head and neck cancers.<br>**Patients can have multiple affected nerves; CN XII and XI EMG confirmed; CN IX/X based on clinical exam features<br>*** LCNP nerve laterality in relation to primary tumor |  |  |  |

### Feasibility and tolerability

In the Phase I trial participants, a total of eight adverse events were attributed to the study drug. Insomnia (n = 4) was the most frequently reported event. All insomnia events were CTCAE grade 1 and considered definitely related. Insomnia started during the corticosteroid administration period and, in several cases, persisted into the tapering phase. One patient in the Dose 2 group required pharmacologic management. One Dose 2 patient experienced grade 1 worsening of previously stable depression during the tapering phase, which was likely related to the study drug. Another patient developed grade 3 hypertension that was present prior to corticosteroid initiation but worsened during the high-dose and tapering phases and was conservatively managed with monitoring alone. No patients in the trial cohort required treatment discontinuation or dose modification due to attributed AE. Both dose regimens were deemed feasible and tolerable per the pre-specified criteria.

The only additional attributable AE identified in the registry cohort was a single case of fatigue occurred within 1–2 weeks post-taper and was considered possibly related. No grade ≥2 toxicities beyond insomnia were observed, and no patients required treatment modification or drug discontinuation.

### Response assessment in Phase I Trial Participants

#### MDASI-HN Top-5 Mean Score Results

At 1–2 weeks post-taper, the median MDASI-HN Top-5 mean score change was -0.2 (IQR: 1.2) for Dose 1 and -1.6 (IQR: 3.6) for Dose 2 (Supplementary Table 3, Figure 2). By 6–10 weeks post-taper, the median change in Dose 1 was -0.8 (IQR: 2.0), while in Dose 2, it was 0 (IQR: 1.85). Three patients in the Dose 2 cohort met the predefined improvement threshold (≥ -1.315 units) at 1-2 weeks post-taper, as indicated by the waterfall plot (Figure 2a) but none met the threshold at 6-10 weeks. In contrast, only one patient in Dose 1 met the threshold at either timepoint (Figure 2b). Per protocol, the pre-specified criteria for Phase II progression were met at Dose 2, and the trial will advance to Phase II at this dose level.

**Figure 2:**
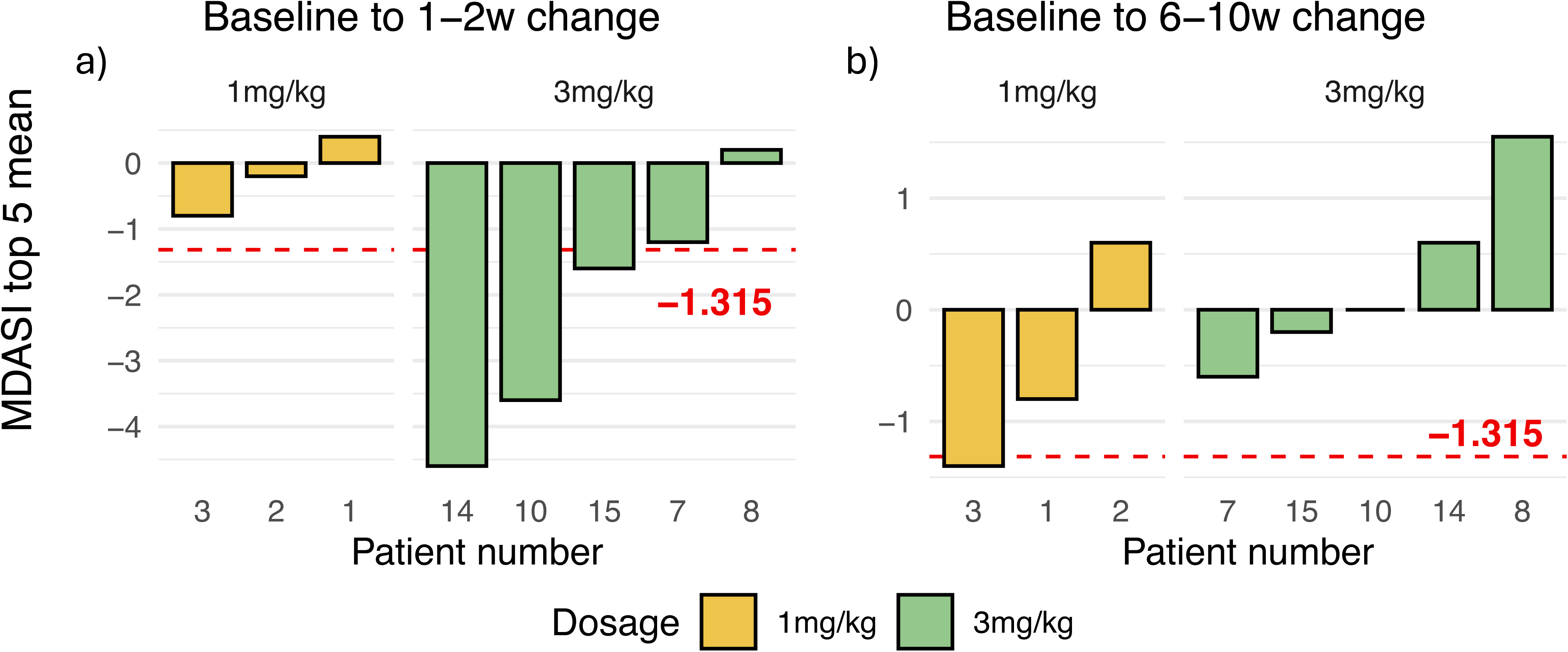
Waterfall plots showing change in MDASI scores from baseline following corticosteroid taper. Panels a and b represent change in MDASI Top 5 mean scores at 1-2 weeks and 6-10 weeks post-taper, respectively. Negative scores indicate improvement. Gold and green bars correspond to the Dose 1 (1 mg/kg) and Dose 2 (3 mg/kg) cohorts, respectively. The dashed red line, shown only in panels a and b, denotes the predefined threshold for clinically meaningful improvement (≥ - 1.315 units).

Per one sample t-test. In the Dose 1 cohort (1 mg/kg), the mean symptom change was −0.96 (95% CI: −2.41 to 0.50; p = 0.17). In the Dose 2 cohort (3 mg/kg), the mean change was −2.16 (95% CI: −4.55 to 0.23; p = 0.066). Although not powered for formal significance testing, these comparisons support a potential dose-response relationship.

#### Patterns of individual symptom response in Phase I Participants

Inspecting single MDASI-HN symptom changes underlying the top 5 mean change (Supplementary Figures 2-3) suggests this is mostly driven by early improvements in bulbar symptoms (voice/speech, swallow/chew, choke) along with mucus symptom reduction in Dose 2 and later improvements in Dose 1, largest in a single patient for mucus.

#### Secondary Endpoint Assessment in Phase I Trial Participants

Despite short-term PRO improvement demonstrated on top 5 MDASI-HN scores, neither Dose I nor Dose II demonstrated consistent patterns of meaningful improvement in clinician-graded functional outcomes (Table 2). At baseline, Dose 1 patients demonstrated worse scores in most functional and patient-reported measures, including MDADI, PSS-HN diet, lingual strength, range of motion, and feeding tube dependence, compared to the Dose 2 group. In the Dose 1 group, MDADI scores improved modestly at 1–2 weeks before declining by 6–10 weeks. PSS-HN diet scores followed a similar pattern, and lingual strength and L-ROM showed transient gains followed by stability or decline. In contrast, Dose 2 patients had relatively stable scores at 1–2 weeks with more observable improvements by 6–10 weeks across all metrics. L-ROM increased steadily from baseline to 6–10 weeks. Missing data, including the patient with the worst clinician-graded function at baseline not returning for the final timepoint, limits interpretation of these trends. Feeding tube status remained unchanged in both Dose levels. Clinician-rated swallowing function using the Dynamic Imaging Grade of Swallowing Toxicity (DIGEST) did not demonstrate consistent improvement across dose levels and is reported in the Supplementary Materials.

**Table 2:**
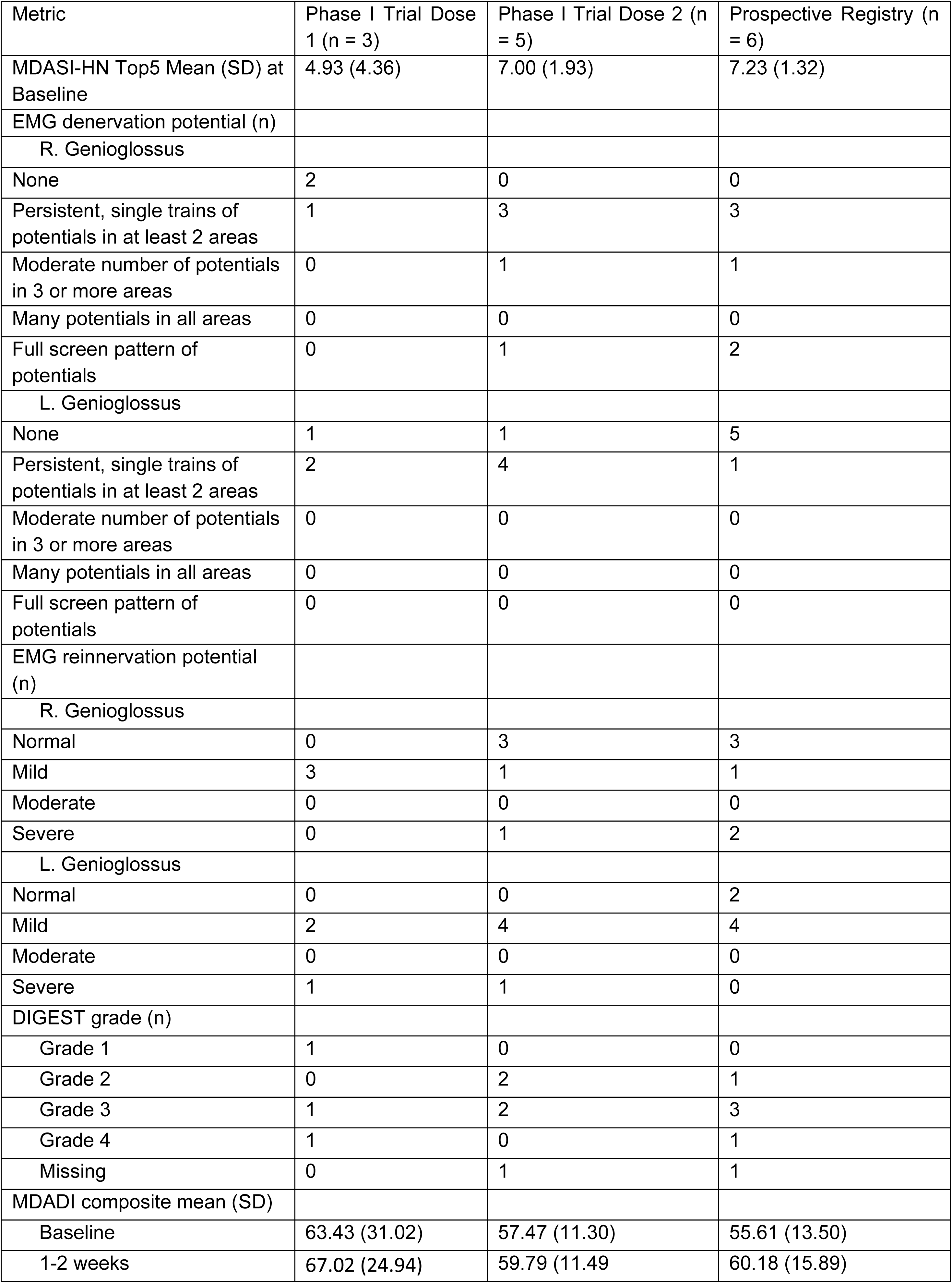
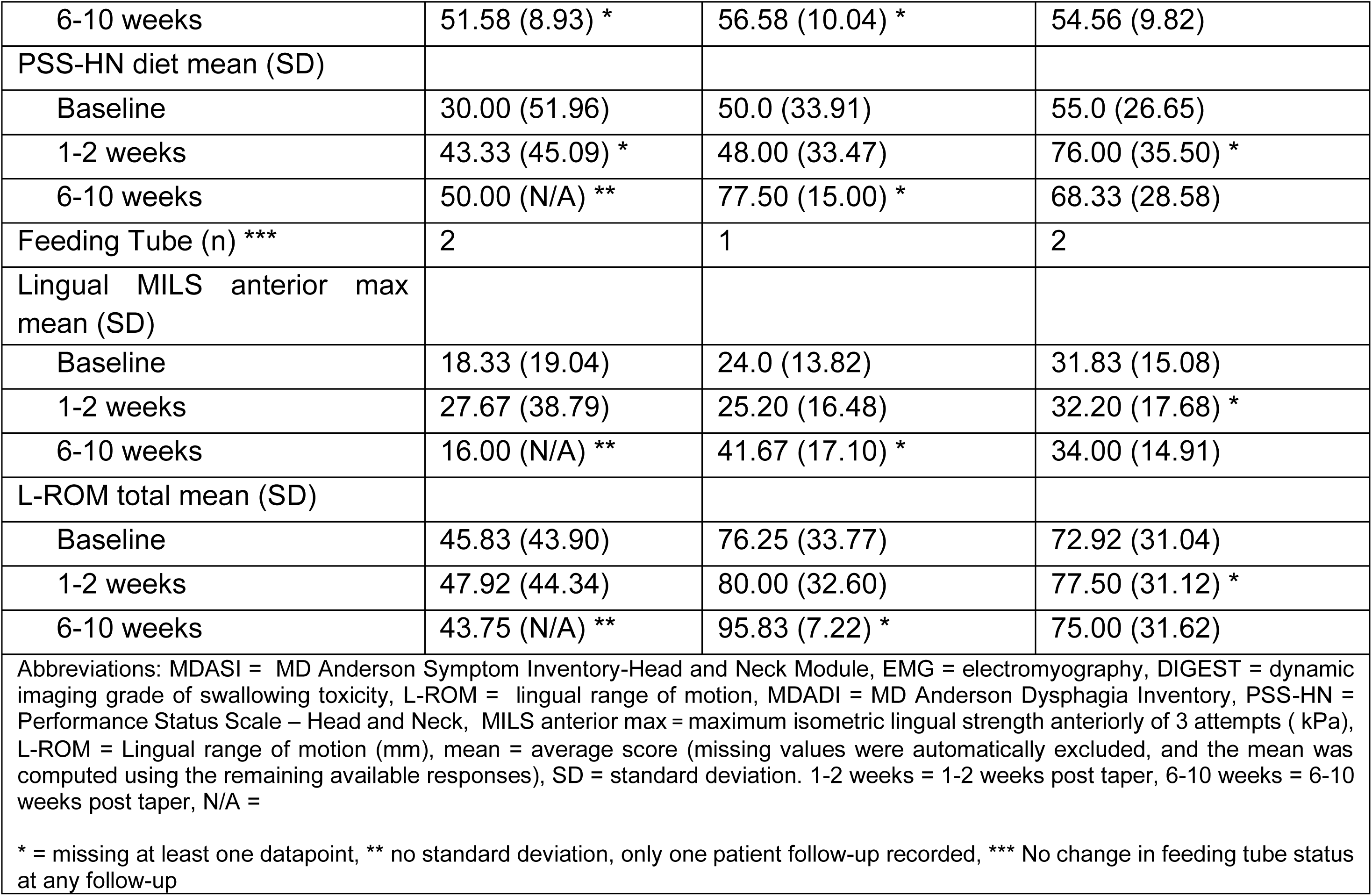
Baseline function and secondary endpoints of clinician-graded function and patient-reported outcomes.

#### Electrophysiological Assessments

For Dose 1 (n=3), denervation responses at 1–2 weeks post-taper were classified as improved in one patient and mixed in two. Reinnervation responses were stable in one patient, improved in one, and mixed in another. At 6–10 weeks, one patient showed mixed denervation and reinnervation responses, and two had no follow-up data.

In the Dose 2 group (n=5), at 1–2 weeks post-taper, three patients demonstrated stable denervation and reinnervation responses. One patient showed improved denervation and mixed reinnervation, while another had no data available at this timepoint. By 6–10 weeks, only two patients had follow-up data, both of whom remained stable; the remaining three lacked scores at this interval. No patients in this group showed worsening in either domain at either interval.

#### Exploratory 2-year post-taper MDASI symptom trajectories by group

Figure 3 shows individual patient-level trajectories of mean Top 5 MDASI symptom scores from baseline through 24 months in Phase I trial participants, stratified by corticosteroid dose. Symptom trajectories were variable between 1–2 weeks and 12 months post-taper in both cohorts. By 18–24 months, the 3 mg/kg group demonstrated a mixed response: 2 of 5 patients showed improvement in mean scores, 2 worsened, and 1 remained stable. A similar pattern was observed in the 1 mg/kg group, where 1 patient improved, 1 worsened and 1 remained stable.

**Figure 3:**
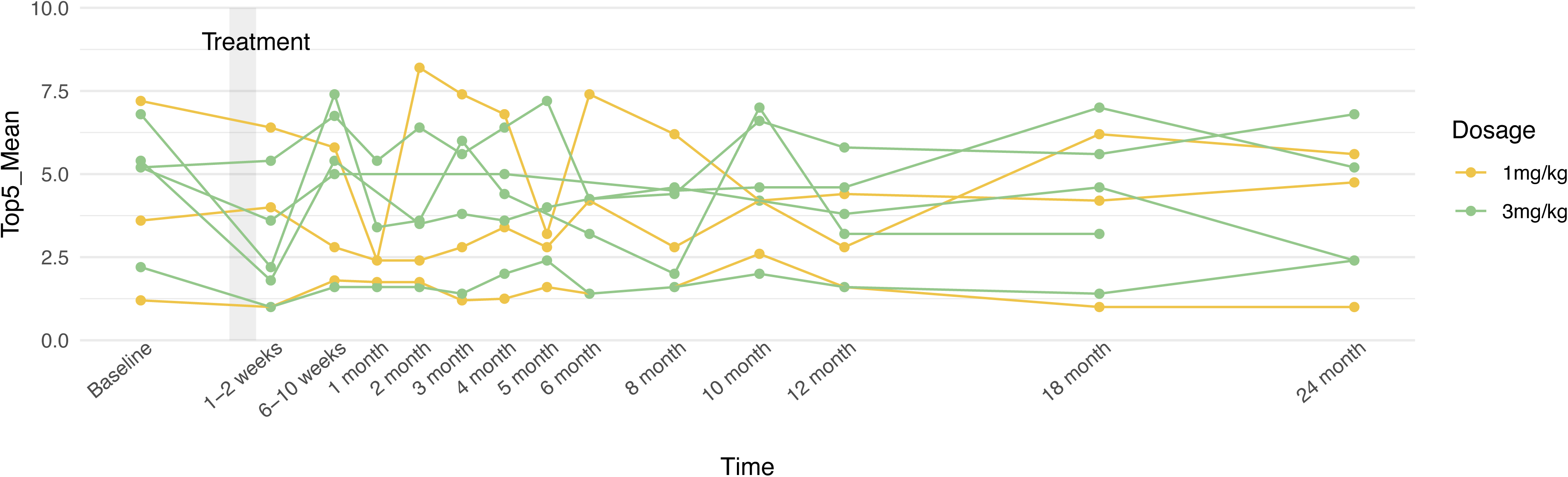
Spaghetti plots for each trial patient per follow up from baseline to 2 years post-taper. Yellow and green represent MDASI Top 5 mean scores for the Dose 1 (1 mg/kg) and Dose 2 (3 mg/kg) cohort respectively. The vertical grey band shows the corticosteroid administration period.

### Prospective Registry (Exploratory)

A similar analysis was performed on the prospective registry patients and can be seen in Supplementary Materials.

## Discussion

Cranial neuropathy is a progressive, potentially debilitating latent sequelae of head and neck radiotherapy with no proven therapies. This prospective Phase I dose-finding trial evaluated the feasibility, tolerability, and preliminary efficacy of high-dose corticosteroids in patients with radiation-associated lower cranial neuropathy (LCNP). Both prednisone dose levels (1 mg/kg and 3 mg/kg) were feasible and well tolerated, with no treatment discontinuations, or dose modifications, and only one significant adverse event that resolved with observation. The primary response endpoint, defined by improvement in the MDASI-HN top 5 mean score, demonstrated early symptomatic benefit in the Dose 2 cohort, with 3 of 5 patients meeting the pre-specified clinically meaningful improvement threshold at 1–2 weeks post-taper. No additional responders were observed at 6–10 weeks, and Dose 2 (3mg/kg) met criteria to move forward to Phase II testing.

Dose 1 (1 mg/kg) did not meet the response threshold at either timepoint. This dose was previously reported to results in measurable neurophysiologic and functional improvements in a single case report [15] that were not replicated in our Phase I testing. While our Phase I symptom trends suggest a dose and time-dependent response, the study design did not afford statistical power for inferential statistical testing

For a progressive, late effect of therapy a response window of 10 weeks is not the target. The goal is durable symptomatic improvement or halting the progressive deterioration in symptoms. Exploratory 2-year follow-up data revealed heterogeneous but potentially durable symptom responses within a subset of patients in both dose cohorts that merits further study. It will be essential to clarify the durability of response and better define which patients are most likely to benefit from corticosteroid therapy in later phase studies.

Beyond PROs, some of the additional patient-reported secondary endpoints as well as tongue specific measures —including PSS-HN, MDADI, MILS, and L-ROM—showed trends generally concordant with MDASI-HN across cohorts, particularly in the Dose 2 and Registry groups. These exploratory findings suggest potential for delayed improvement in functional domains but remain hypothesis-generating due to small sample size and incomplete data. In addition to biologic or treatment-related mechanisms, potential contributors to delayed improvement may include psychosocial or behavioral factors such as the Hawthorne effect, response shift, or hedonic adaptation, all of which can influence patient perception and reporting over time [36,37,38].

While patient-reported and functional outcomes showed potential improvements that encourage further testing, more objective measures such as EMG and DIGEST did not demonstrate consistent recovery across cohorts. These assessments largely reflected stable or mixed neuromuscular findings with minimal short-term change, with interpretation challenged like other measures by missing values. These findings are consistent with prior reports showing limited early recovery in swallowing physiology following radiation-associated cranial neuropathy [13,35,39], and highlight the often reported discordance between subjective symptom improvement and clinician-graded neuromuscular recovery.

These findings align with clinical experience and the limited literature indicating that radiation-induced cranial neuropathies result in chronic neuromuscular deficits that are difficult to measurably improve on imaging based measures like DIGEST [40]. Thus, an important limitation of the response assessment is our reliance on a PRO. While the **** Symptom Inventory–Head and Neck Module (MDASI-HN) is a validated tool for symptom assessment [32], prior research has shown that patient-reported symptom scores do not always correlate with objective functional outcomes, reflecting the complex relationship between symptom perception and functional recovery in this population [10,32,39].

This study benefits from a prospective Phase I/II design testing a well-tolerated drug for a debilitating orphan indication with no proven therapies. A robust patient testing protocol blending gold-standard needle EMG, validated clinician-graded measures of tongue and swallow function, and a host of PROs also affords comprehensive insights to generate hypotheses for next phases of this work. The trial, however, has several limitations. As a Phase I trial, the small sample size limits power to detect statistical differences or draw definitive conclusions about clinical efficacy. Although corticosteroids are regularly used in neurologic conditions, the go forward dosing (3 mg/kg) was pragmatically designed and has not been previously tested for this indication, limiting our ability to compare or interpretation observed responses in context of prior work. Missing data across all groups for secondary endpoints further introduces important constraints when interpreting results in small subgroups of patients. The Registry design also introduced variability in data capture, inclusion criteria, and baseline functional status, including broader tumor types and surgical histories. Finally, while validated, the MDASI-HN reflects symptom burden rather than functional capacity and may diverge from objective recovery measures. Thus, symptom responses meeting criteria for Phase II testing may not reflect meaningful physical changes in function.

### Conclusions

These results suggest that corticosteroids may offer short-term symptomatic benefit in patients with radiation-associated LCNP, particularly at higher doses. Even transient relief may be clinically meaningful in this population, where swallowing dysfunction, choking, and voice changes significantly affect quality of life and nutritional independence. Exploratory functional outcomes suggest potential for delayed improvement, particularly at higher corticosteroid doses, though this requires confirmation in larger studies. A Phase II trial with greater statistical power, structured functional endpoints, and extended follow-up will be critical to determine whether early symptom benefits translate into durable, clinically meaningful functional improvements. 2-year MDASI-HN data showed no significant trends but further analysis in a larger phase II cohort could aid in revealing subgroups where corticosteroid treatment can provide long-term benefits.

## Data Availability

In accordance with NOT-OD-21-013, Final NIH Policy for Data Management and Sharing, anonymized/de-identified data that support the findings of this study are openly available in an NIH-supported generalist scientific data repository (figshare) at 10.6084/m9.figshare.31082410 no later than the time of an associated publication in a peer reviewed journal.

https://doi.org/10.6084/m9.figshare.31082410

## Funding Acknowledgement

This work was supported directly or in part by funding/resources from the National Institutes of Health (NIH) National Cancer Institute (1P01CA285249); the MD Anderson Cancer Center Support Grant Biostatistics Resource Group (P30CA016672), the Charles and Daneen Stiefel MD Anderson Oropharynx Fund and HESI/Thrive Foundation funding. SYL and CDF received related funding for data collection from NIH National Institute of Dental and Craniofacial Research (NIDCR; R01DE025248, R01DE028290); SYL, KAH, and CDF received related technical development support from NCI (R01CA218148). CDF received relevant funding for data analysis from a joint National Science Foundation (NSF)/NCI award through NCI (R01CA257814).

## Data sharing statement

In accordance with NOT-OD-21-013, Final NIH Policy for Data Management and Sharing, anonymized/de-identified data that support the findings of this study are openly available in an NIH-supported generalist scientific data repository (figshare) at 10.6084/m9.figshare.31082410 no later than the time of an associated publication.

## Public access policy compliance

In accordance with NOT-OD-25-049, Supplemental Guidance to the 2024 NIH Public Access Policy: Government Use License and Rights,: “This manuscript is the result of funding in whole or in part by the National Institutes of Health (NIH). It is subject to the NIH Public Access Policy. Through acceptance of this federal funding, NIH has been given a right to make this manuscript publicly available in PubMed Central upon the Official Date of Publication, as defined by NIH.”

## CRediT statement

In accordance with the Contributor Roles Taxonomy (CRediT, https://credit.niso.org/), the contributing authors have designated responsibilities and individual author attribution. The corresponding author(s) (KAH) assume(s) responsibility for role assignment, and all contributors have been given the opportunity to review and confirm assigned roles.

- ZB: Methodology, Formal analysis, Investigation, Data curation, Visualization, Writing, Review & Editing
- CBP: Methodology, Software, Validation, Formal analysis, Writing, Review & Editing
- CEB: Data curation, Investigation, Review & Editing
- HM: Data curation, Review & Editing
- JAG: Data curation, Review & Editing
- SB: Investigation, Data curation, Review & Editing
- KW: Investigation, Methodology, Data curation, Review & Editing
- CDF: Resources, Funding acquisition, Review & Editing
- SYL: Conceptualization, Funding acquisition, Supervision, Project administration, Review & Editing, Corresponding Author
- KAH: Conceptualization, Funding acquisition, Methodology, Supervision, Project administration, Writing, Review & Editing, Corresponding Author

## Declaration of generative AI and AI-assisted technologies in the manuscript preparation process

During the preparation of this work, the author(s) used ChatGPT (OpenAI, San Francisco, CA) to assist in language refinement, structural organization, and formatting of the manuscript. The author(s) reviewed and edited the content as needed and take full responsibility for the accuracy and integrity of the final manuscript.

## Supplementary Figure and Table Descriptions

**Supplementary Figure 1:**
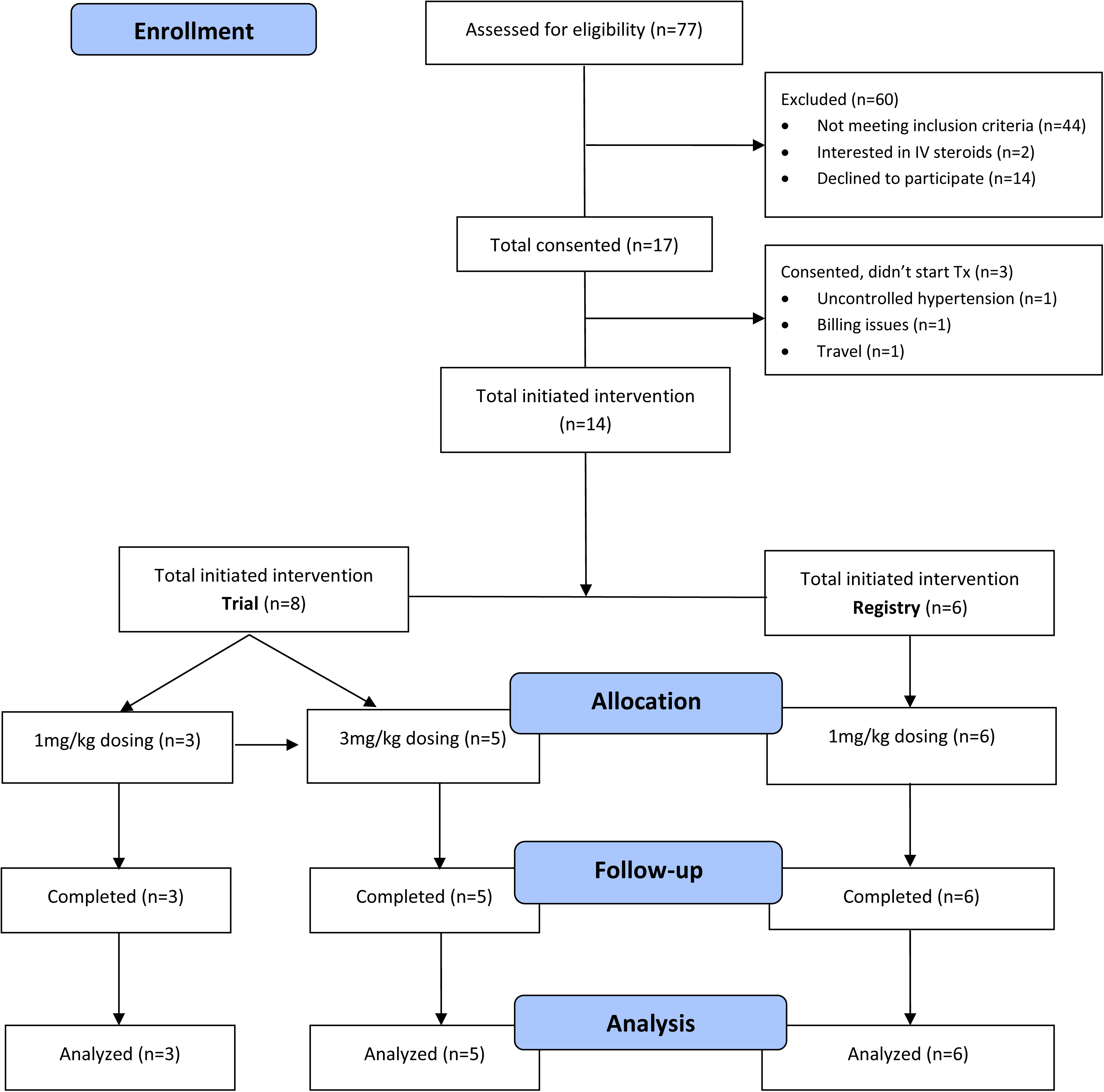
CONSORT diagram used in the XXXX trial.

**Supplementary Table 1:** Full Trial and Registry Eligibility Criteria.

| Timepoint, Arm | Median | Standard Dev | Quartile 1 | Quartile 3 | Interquartile Range |
| --- | --- | --- | --- | --- | --- |
| 1-2 weeks, Dose 1 | -0.2 | 0.6 | -0.8 | 0.4 | 1.2 |
| 6-10 weeks, Dose 1 | -0.8 | 1.0 | -1.4 | 0.6 | 2 |
| 1-2 weeks, Dose 2 | **-1.6** | 1.9 | -4.1 | -0.5 | 3.6 |
| 6-10 weeks, Dose 2 | 0 | 1.0 | -0.78 | 1.08 | 1.85 |
| 1-2 weeks, Registry | -0.2 | 1.2 | -1.4 | 1.45 | 2.85 |
| 6-10 weeks, Registry | **-1.5** | 2.0 | -2.85 | 1.49 | 4.34 |
| Abbreviations: 1-2 weeks = 1-2 weeks post-taper, 6-10 weeks = 6-10 weeks post-taper, Dose 1 & Registry = 1mg/kg, Dose 2 = 3 mg/kg<br>**\*Cutoff for clinical significance is –1.315 denoted in bold.** |  |  |  |  |  |

**Supplementary Table 2:** Schedule of Assessments.

| Assessment | Method | Domain | Endpoint(s) | Scale | Pre-ST | Post-Taper<sup>a</sup> | 6-10 weeks<sup>b</sup> |
| --- | --- | --- | --- | --- | --- | --- | --- |
| **Clinical studies** |  |  |  |  |  |  |  |
| EMG (Tongue) | Electromyography | Neuropathy | 4-point denervation potentials grade<sup>26,27,28</sup> | Ordinal | X | X | X |
| MBS | Fluoroscopy | Dysphagia severity grade | DIGEST<sup>29,30</sup> | Ordinal: 0 (best), 4 (worst) | X | X | X |
| Lingual function | Lingual ROM and manometric pressures | Tongue strength and ROM | Maximum isometric lingual strength (MILS); L-ROM<sup>31</sup> | Continuous; Ordinal | X | X | X |
| **Questionnaires** |  |  |  |  |  |  |  |
| MDASI-HN<sup>32</sup> | PRO (28-item) | Symptom burden | Symptom severity, symptom interference | Continuous: 0 (best), 10 (worst) | X | X | X<sup>c</sup> |
| MDADI<sup>33</sup> | PRO (20-item) | Swallowing-related QOL | Composite, Global, & subscale scores | Continuous: 20 (worst), 100 (best) | X | X | X |
| PSS-HN<sup>34</sup> | Interview (3-item) | Functional status | Diet, Eating, Speech subscales | Ordinal: 0 (worst), 100 (best) | X | X | X |

Abbreviations: ST, corticosteroid therapy, EMG, electromyography, MBS, modified barium swallow, DIGEST, dynamic imaging grade of swallowing toxicity, ROM, range of motion, L-ROM, lingual range of motion, MDASI-HN, MD Anderson Symptom Inventory-Head and Neck Module, PRO, patient-reported outcome, QOL, Quality of Life, MDADI, MD Anderson Dysphagia Inventory, PSS-HN, Performance Status Scale – Head and Neck
<sup>a</sup>1-2 weeks post-taper; <sup>b</sup> 6-10 weeks post-taper;
<sup>c</sup>Collected through 2-years post therapy
Both shaded and unshaded timepoints were collected on clinical trial participants, while a more limited set of data in the shaded cells were completed by patients on the registry. All unshaded timepoints were optional to the registry patients or were completed by clinician's discretion.

**Supplementary Table 3:** MDASI Top-5 Mean score analysis by median, standard deviation and interquartile range.

| **Inclusion Criteria** | **Clinical Trial** | **Registry** |
| --- | --- | --- |
| **Cancer Type** | Oropharyngeal Cancer Only | Any Head and Neck Cancer |
| **Time Since Treatment** | ≥2 years post-radiotherapy |  |
| **Cranial Nerve Involvement** | EMG confirmed XII ± X |  |
| **Structural/Malignant Source** | Excluded (confirmed by imaging, exam, or biopsy) |  |
| **Follow-Up** | Required (must return for in person post-steroid therapy assessment) | Optional in person follow-up testing |
| **Language** | Required (English, Simplified Chinese, Filipino, Greek, Japanese, Korean, Russian, Spanish, Taiwanese) |  |
| **Prior Surgery Near Hypoglossal Nerve Path** | Excluded | Included |
| **Uncontrolled Diabetes** | Excluded |  |
| **Uncontrolled Hypertension (systolic >160, diastolic >90)** | Excluded |  |
| **Gastrointestinal Ulcer** | Excluded |  |
| **History of Psychosis** | Excluded |  |
| **Bipolar Disorder** | Excluded |  |
| **Obstructive Pharyngoesophageal Stricture** | Excluded |  |
| **Implanted Pacemaker or Electrical Device** | Excluded |  |
| **Pregnancy** | Excluded |  |

**Supplementary Figure 2:**
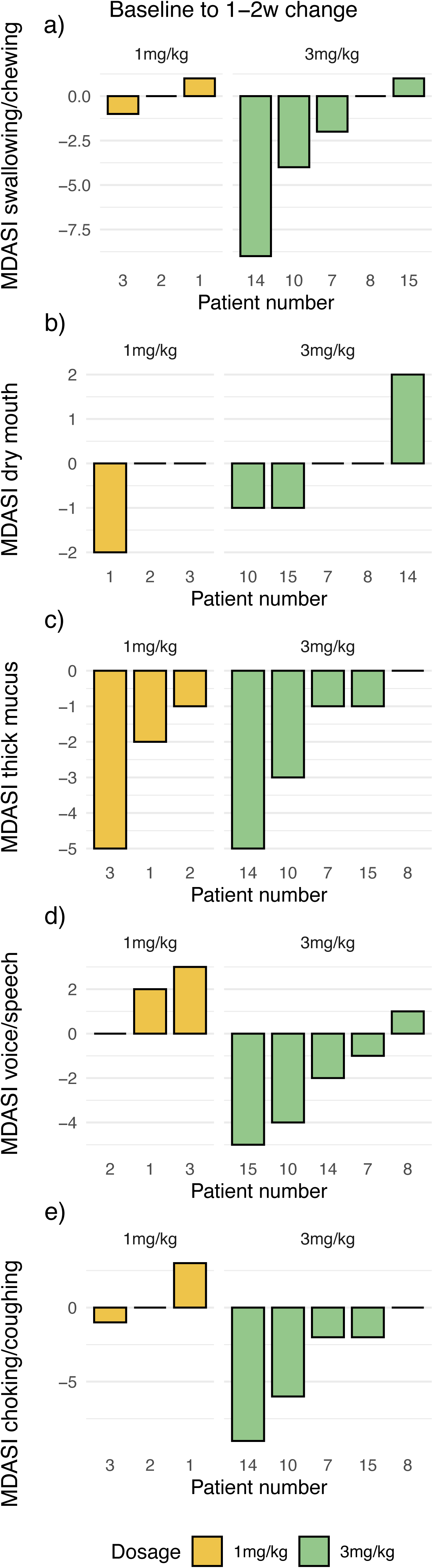
Waterfall plots seen in panels a - e represent change in individual patient MDASI symptom scores from baseline to 1-2 weeks (1-2w) post corticosteroid taper. Negative scores indicate improvement. Gold and green bars correspond to the Dose 1 (1 mg/kg) and Dose 2 (3 mg/kg) cohorts, respectively.

**Supplementary Figure 3:**
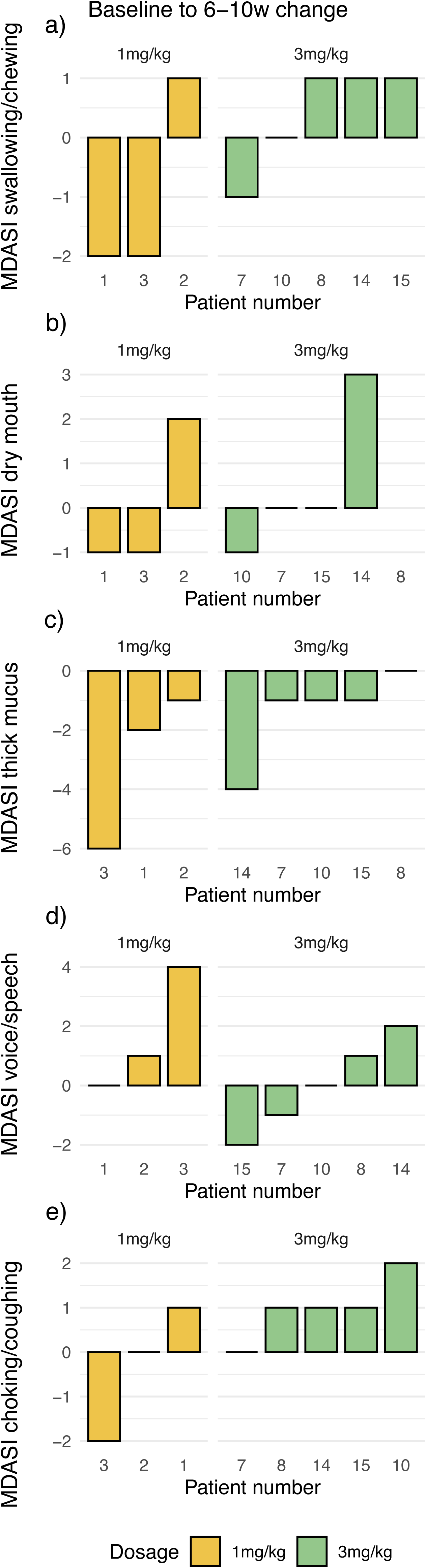
Waterfall plots seen in panels a - e represent change in individual patient MDASI symptom scores from baseline to 6-10 weeks (6-10w) post corticosteroid taper. Negative scores indicate improvement. Gold and green bars correspond to the Dose 1 (1 mg/kg) and Dose 2 (3 mg/kg) cohorts, respectively.

**Supplementary Table 4:**
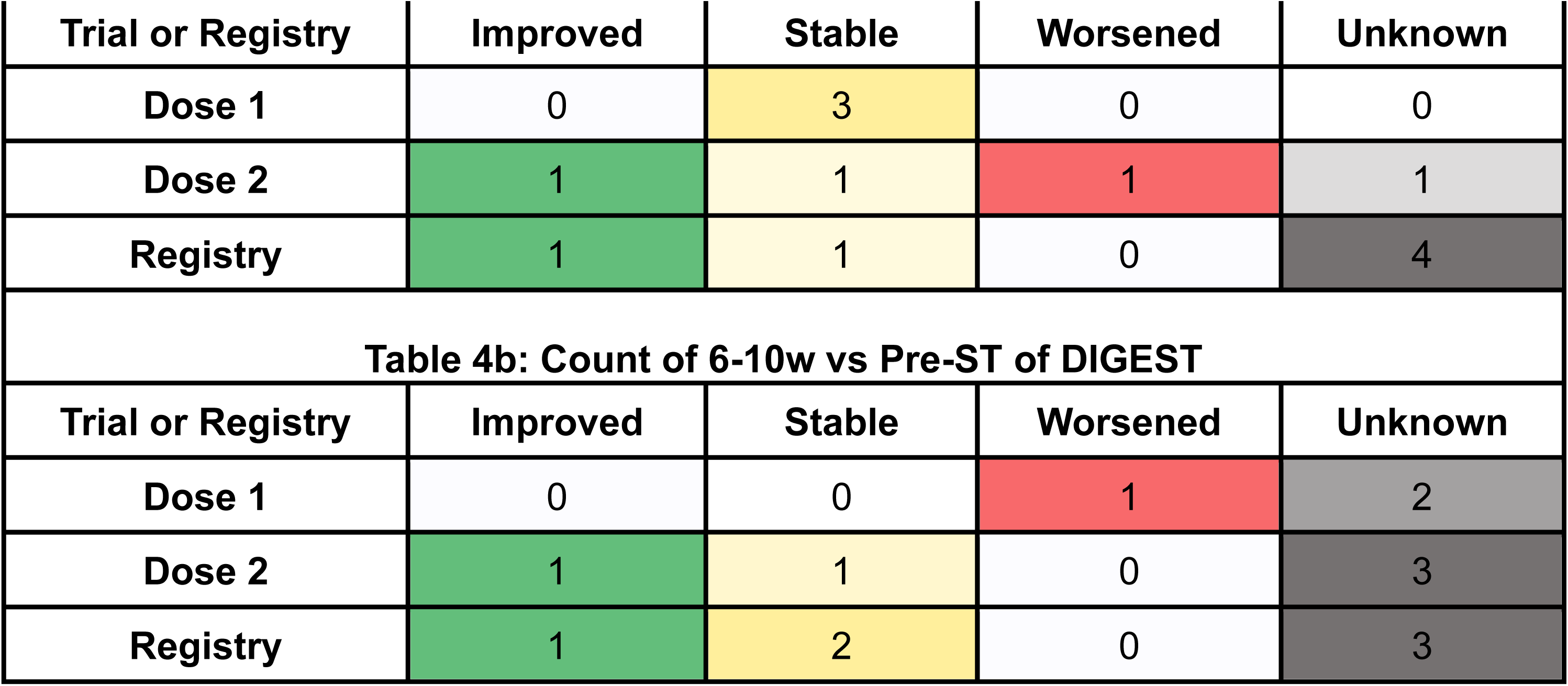
Contingency table representing the change in DIGEST scores from Baseline to 1-2 weeks post-taper (Table 4a, top) and Baseline to 6-10 weeks post-taper (Table 4b, bottom). Cell shading denotes DIGEST change from pre-steroid baseline: green = improved, yellow = stable, red = worsened, gray = unknown.

### DIGEST Review

Among patients in the 2 trial cohorts, clinician-graded image based DIGEST scores showed minimal early improvement following steroid therapy (Supplementary Table 4). At 1–2 weeks post-taper, only one patient in the Dose 2 group demonstrated improvement relative to baseline, while most patients in both trial arms had stable function (Supplementary Table 4a). By 6–10 weeks, two patients—one from each trial arm—had improved swallowing function. Worsening was uncommon across both timepoints, and a notable proportion of patients, particularly in the Dose 2 group at 6–10 weeks, had missing data classified as unknown (Supplementary Table 4b).

### Prospective Registry (Exploratory)

#### MDASI-HN Top-5 Mean Score Results

The registry group received 1 mg/kg prednisone (same as Dose 1) and demonstrated a median MDASI-HN Top-5 mean score change of -0.2 (IQR: 2.85) at 1-2 weeks post-taper and -1.5 (IQR: 4.34) at 6-10 weeks post-taper (Supplementary Table 3). Two patients in the registry group met the improvement threshold at 1-2 weeks, and three patients met the threshold at 6-10 weeks (Supplementary Figures 4a and 4b). While registry participants exhibited symptom improvement, their outcomes do not contribute to Phase II dose selection due to differing inclusion criteria and the exploratory nature of the registry.

In the registry cohort (Supplementary Figure 4a and 4b), delayed symptom improvement was more prominent. Earlier improvements at 1–2 weeks were observed in 3 patients for voice, 4 for mucus and dry mouth, and 2 for swallowing and choking (Supplementary Figure 5). At 6–10 weeks, 5 of 6 patients improved in voice and swallowing, 4 in mucus, and 3 in choking and dry mouth (Supplementary Figure 6).

#### Functional Measures Over Time

Baseline functional measures for the prospective registry cohort are summarized in Table 2. MDASI-HN Top 5 scores were highest in this group (mean 7.2 ± 1.3). EMG showed right-sided denervation in all patients that was generally low-grade, though two exhibited a denervation score of 4. Left-sided denervation was less prominent. Reinnervation scores were typically mild, with two patients demonstrating right-sided scores of 4. DIGEST grade 3 or higher was observed in three of six patients.

Additional swallowing-related patient-reported and functional outcomes over time are shown in Table 2. At baseline, registry patients had scores comparable to the Dose 2 group across MDADI, PSS-HN diet, lingual strength, and range of motion. MDADI scores improved modestly at 1–2 weeks (60.2) before declining slightly by 6–10 weeks (54.6), with complete follow-up available for all patients. PSS-HN diet scores increased steadily over time (55.0 → 76.0 → 68.3), with upward trends also observed in lingual strength and L-ROM, particularly at later timepoints. At least one patient had missing 1–2-week data for PSS-HN, MILS, and L-ROM; however, all patients had complete follow-up data at 6–10 weeks. Feeding tube status remained unchanged in the two dependent patients.

#### Electrophysiological & Swallowing Assessments

A qualitative review of bilateral genioglossus denervation and reinnervation changes was performed for the Registry group in a similar fashion to the trial groups. Denervation responses at 1–2 weeks were stable in two patients, mixed in three, and not available in one. Reinnervation responses were stable in four patients, mixed in one, and not available in one. These patterns remained consistent at 6–10 weeks, with no additional cases of worsening or improvement.

#### DIGEST Review

In the Registry cohort, early changes in DIGEST scores were similarly limited, with one patient showing improvement and one stable function at 1–2 weeks post-taper, while the majority had unknown outcomes (Supplementary Table 4a). By 6–10 weeks, the number of patients had stable or improved swallowing function increased slightly, with no cases worsening. However, the overall interpretability of the registry data was limited by the high proportion of missing follow-up assessments (Supplementary Table 4b).

#### 2 year MDASI symptom trajectories

Supplementary Figure 7 shows individual patient-level trajectories of mean Top 5 MDASI symptom scores from baseline through 24 months in the registry cohort. Symptom trajectories were variable during the first 10 months post-taper. By 24 months, 3 of 6 patients demonstrated improvements in mean MDASI scores compared to baseline, 2 patients were stable, and 1 additional patient was lost to follow-up after 4 months.

## Supplementary Figure and Table Descriptions

**Supplement Figure 4:**
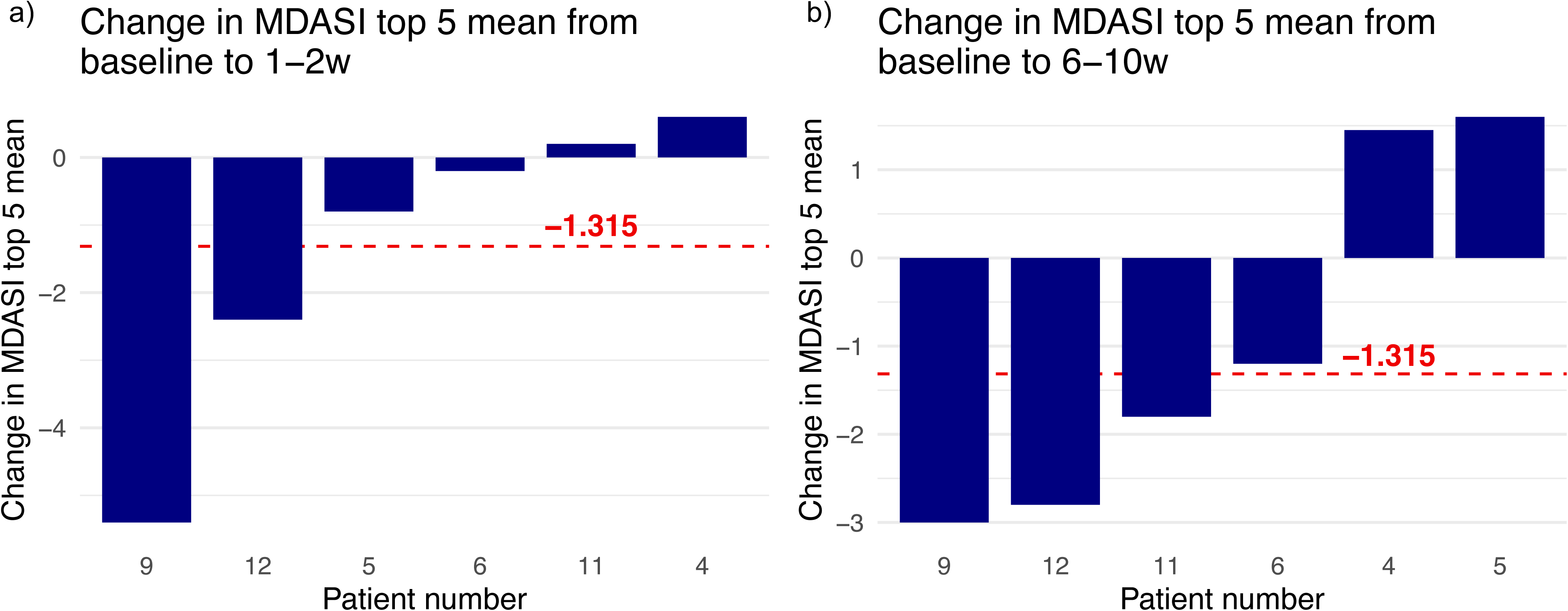
Waterfall plots represent the Registry cohort (1 mg/kg) change in MDASI top 5 mean scores from baseline to 1-2 weeks (Figure 4a, left) and 6-10 weeks (Figure 4b, right) post-taper with negative scores indicating improvement. The dashed red line shows the predetermined improvement threshold (≥ -1.315 units).

**Supplementary Figure 5:**
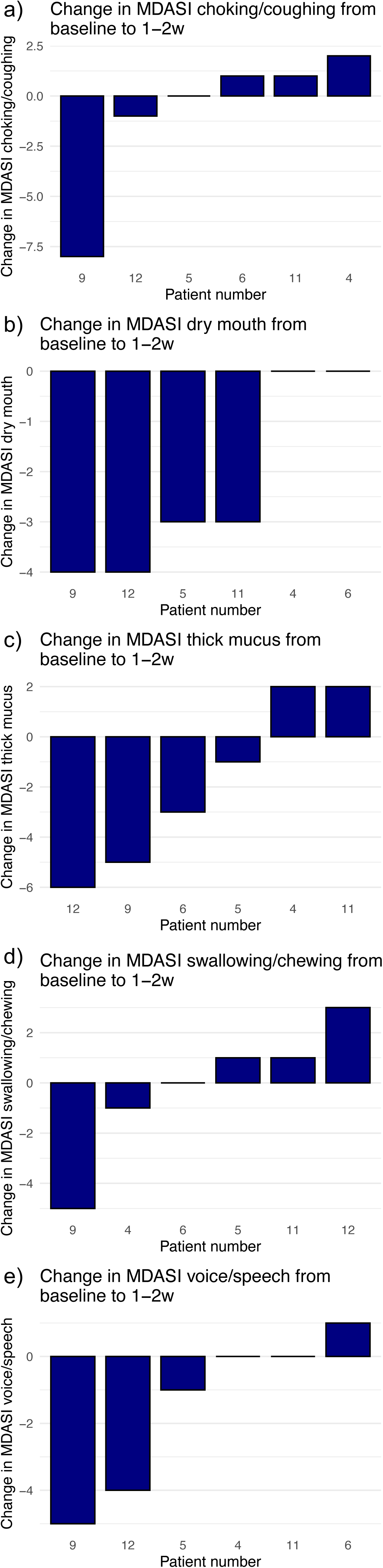
Waterfall plots (a-e) represent the change in individual MDASI scores of the Registry cohort (1 mg/kg) per symptom from baseline to 1-2 weeks post-taper with negative scores indicating improvement.

**Supplementary Figure 6:**
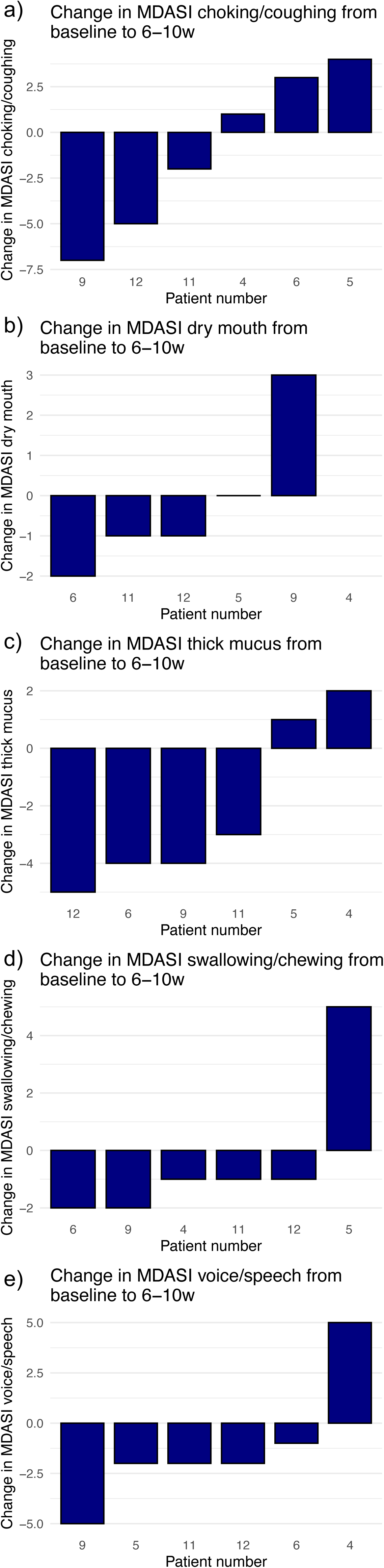
Waterfall plots (a-e) represent the change in individual MDASI scores of the Registry cohort (1 mg/kg) per symptom from baseline to 6-10 weeks post-taper with negative scores indicating improvement.

**Supplementary Figure 7:**
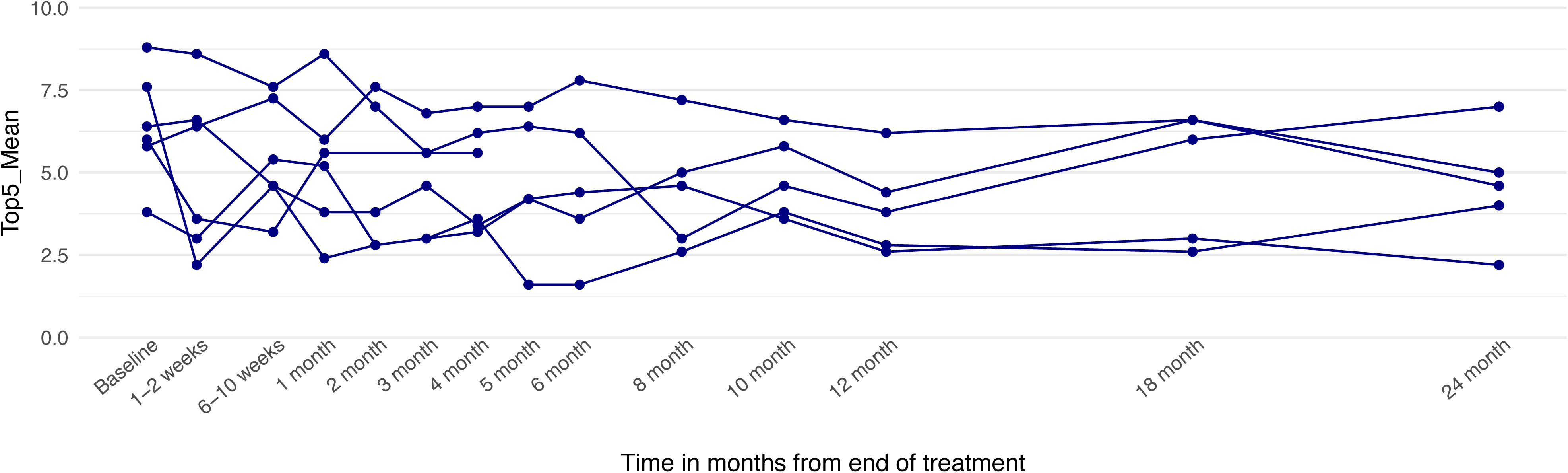
Spaghetti plots for each trial patient per follow up from baseline to 2 years post-taper. Blue represents MDASI Top 5 mean scores for the Registry cohort (1 mg/kg).

